# CT-Perfusion absolute Ghost Infarct Core is a rare phenomenon associated with poor collateral status in acute ischemic stroke patients

**DOI:** 10.1101/2024.07.09.24310115

**Authors:** Giorgio Busto, Andrea Morotti, Ilaria Casetta, Anna Poggesi, Davide Gadda, Andrea Ginestroni, Giorgio Arcara, Arianna Rustici, Andrea Zini, Alessandro Padovani, Enrico Fainardi

## Abstract

**Background:** CT-Perfusion (CTP) overestimation of core volume >10 mL compared to the final infarct volume (FIV) size is the current definition of the ghost infarct core (GIC) phenomenon. However, subsequent infarct growth might influence FIV. We aimed to report a more reliable assessment of GIC occurrence, defined as the lack of FIV at 24 hours follow-up imaging, compared to CTP core volume at admission. This phenomenon was named absolute GIC (aGIC) and we investigated its prevalence and predictors.

**Methods:** A total of 652 consecutive stroke patients with large vessel occlusion who achieved successful recanalization (mTICI 2b-3) after Endovascular Treatment (EVT) and non-contrast CT (NCCT) follow-up imaging at 24 hours were retrospectively analyzed. Ischemic core volume was automatically generated from CTP, and FIV was manually determined on follow-up NCCT. Multivariable logistic regression was used to explore aGIC predictors.

**Results:** We included 652 patients (53.3% female, median age 75 years), of whom 35 (5.3%) had an aGIC. The aGIC group showed higher ASPECTS (p<0.001), shorter (<3 hours) onset-to-imaging time (p<0.001), poorer collaterals (p<0.001), higher hypoperfusion intensity ratio (p=0.001) compared to the non-aGIC group. In multivariate analysis, ASPECTS (odds ratio [OR], 1.87; p<0.001), onset-to-imaging time (OR, 0.99; p=0.013), collateral score (OR, 0.45; p<0.004) and hypoperfusion intensity ratio (OR, 23.2; p<0.001) were independently associated with aGIC.

**Conclusion:** aGIC is a more reliable evaluation of infarct core volume overestimation assessed on admission CTP and represents a rare phenomenon, associated with ultra-early presentation and poor collaterals.

## INTRODUCTION

Endovascular Treatment (EVT) has dramatically changed the natural history of Acute Ischemic Stroke (AIS) patients with large vessel occlusion (LVO), improving clinical outcome by the reperfusion of ischemic tissue at risk of infarction, namely penumbra, and limiting infarct core growth, that is the irreversible damaged neuronal tissue [1–2]. Different methods to define infarct core volume have been proposed based on two different EVT treatment windows [3]. Identifying the core size with perfusion imaging using relative cerebral blood flow (rCBF) is mandatory to assess eligible patients for EVT between 6 and 24 hours from stroke onset [4]. Conversely, in patients with stroke onset up to 6 hours from last known well the use of advanced imaging is not recommend, to avoid delays due to the elaboration of perfusion maps from row data [5]. However, several studies showed that patients undergoing EVT in the early time window selected with advanced imaging achieved better functional outcomes after successful recanalization than those who were not, encouraging the use of perfusion imaging also in early presenters [6,7]. As a results, a precise calculation of core volume at presentation is pivotal to recruit more patients for EVT in late time window and improve clinical outcomes of patients treated in the early window when selected with perfusion imaging [7]. In this context, a possible overestimation of core extension by perfusion software has been described [8]. This condition, defined as ghost infarct core (GIC), was usually considered as >10 ml of core size on admission CTP compared to the final infarct volume (FIV) visualized on non-contrast computed-tomography (NCCT) [9–13] or diffusion-weighted imaging (DWI) [13–16] at 24-48 hours and up to 7 days [17] from stroke onset. Several mechanisms were associated to GIC phenomenon but the presence of a short time from stroke onset to imaging [9–17] and poor collateral flow [12,15–17] are currently believed to be the most important. However, core overestimation differed across the studies for incidence and impact on functional outcome [8,13], and predominantly involved white matter [14,16]. On the other hand, FIV varied according to its topography [18]. In addition, although FIV is affected by brain edema formation in the early [19] and late [20] phases of infarct evolution, a significant number of AIS patients demonstrated a late infarct growth beyond 24 hours even after successful EVT [21,22] suggesting that 24-hours ischemic lesion volume measurement could not reliably estimate FIV [23,24]. Therefore, as patient with no early infarct growth commonly do not show late lesion expansion [23], a more reliable assessment of GIC could be obtained when a core volume visible on CTP is associated with the absence of infarcted tissue on 24 hours follow-up imaging. We named this profile as absolute ghost infarct core (aGIC) and aimed to establish its frequency and determinants in a cohort of ischemic stroke patients.

## METHODS

This cohort study was approved by the local ethics board and clinical information were recorded during routine clinical activity (PN OSS_26299). Written informed consent was obtained from each patient or from their legally authorized representatives at admission or waived by the institutional review board. STROBE (Strengthening the Reporting of Observational studies in Epidemiology) guidelines for observational studies were utilized.

### Patient Selection

We retrospectively analyzed a prospectively collected cohort of consecutive patients with AIS with anterior circulation LVO treated with EVT and admitted from January 2017 to September 2023 at Careggi University Hospital of Florence. All patients presenting with suspected AIS with LVO, and no history of renal failure or contrast allergy routinely underwent NCCT, multi-phase CT-angiography (mCTA) of the cervical and intracranial vessels and CTP at admission within 24 hours of symptom onset. Patients were included if they presented at the emergency department with the following criteria: (1) NCCT Alberta Stroke Programme Early Computed Tomography Score (ASPECTS)≥6; (2) diagnosis of AIS within 24 hours from witnessed symptom onset or time last seen well; (3) evidence of internal carotid artery (ICA) and/or middle cerebral artery (MCA) M1 or M2 segment occlusion on CTA; (4) CTP performed at admission; (5) selected for receiving EVT; and (6) follow-up NCCT imaging performed at 24 ± 12 hours. Exclusion criteria were: (1) NCCT ASPECTS<6; (2) age < 18 years; (3) pregnancy; (4) severe pre-stroke disability defined as modified Rankin Scale (mRS)≥4; (5) detection of intracerebral hemorrhage (ICH) on admission NCCT; (6) contraindications to iodinated contrast agent; (7) poor quality of CT acquisition due to motion artifacts; and (8) inability to complete multi-modal CT protocol at baseline and/or 24 hours follow-up NCCT.

### Clinical Assessment

Clinical, demographic and technical data were collected by trained investigators blinded to the outcomes of interest, from the patient’s medical records and a prospectively maintained institutional stroke data-base, including age, sex, pre-stroke functional status (mRS), the presence of stroke risk factors, the interval between symptom onset and neuroimaging, the initial National Institute of Health Stroke Scale (NIHSS) score, the use of intravenous thrombolysis (IVT) and EVT. Clinical outcome was measured using the mRS at 3 months. Good outcome was defined as mRS 0-2 at three months [25].

### Imaging Acquisition

All imaging was conducted on 128-slice scanner (Philips Brilliance iCT, Best, the Netherlands). NCCT helical scans were performed from the skull base to the vertex using these following imaging parameters: 120 kV, 340 mA, 0.6 collimation, 1 second/rotation, and table speed of 15 mm/rotation. 0.7 ml/kg contrast (maximum 90 ml), 5-to 10-second delay from injection to scanning, 120 kV, 251 mAs, 0.75 second/rotation, 0.8/0.4 mm thick slices (imbricated slices), scan time 4 seconds. CTA covered from the carotid bifurcation to vertex. The second and third phase were acquired after a delay of 4 seconds that allows for table repositioning to the skull base. Scanning duration for each additional phase was 3.4 seconds. The axial images were reconstructed at 0.4-mm overlapping sections, and MIP multiplanar reconstructions for axial, coronal, and sagittal images of the circle of Willis were performed with 10 mm thickness at 3mm intervals. CTP studies were obtained with a dynamic first-pass bolus-tracking methodology according to a 2-phase imaging protocol, to avoid the truncation of time density curves, with toggling table technique. The 2-phase acquisition consisted of a first phase every 3.2 seconds for 60 seconds and an additional second phase every 15 seconds for 113 seconds, which started 5 seconds after the automatic injection of 40 ml of non-ionic contrast agent followed by a saline flush of 40 ml at the rate of 4 ml/s. Sections of 8 cm length (across z axis) were acquired at 5 mm slice thickness. The other acquisition parameters 80 kV, 150 mAs, and 0.33 rotation time. All CTP source images were reconstructed with the standard filter and display field of view (DFOV) of 25 cm.

### Imaging Processing and Analysis

The extent of early ischemic changes was evaluated on baseline NCCT using the ASPECTS methodology [26]. mCTA collateral supply was graded by 2 diagnostic neuroradiologists, G.B. with more than 10 years of experience and E.F. with more than 20 years of experience, both blinded to clinical information and CTP outputs on a 6-point scale according to a previously published scoring system [27] in which collaterals were categorized as zero filling in any phase in the affected territory (grade 0), just a few vessels visible in any phase (grade 1), delay of 2 phases and decreased prominence or number of vessels, or delay of 1 phase and some ischemic areas with no vessels (grade 2), delay of 2 phases but the same prominence or number of vessels, or delay of 1 phase with the prominence or number of vessels significantly decreased (grade 3), delay of 1 phase but prominence and extent are the same (grade 4); no delay and normal or increased number or prominence of vessels (grade 5). Grades 0-3 were considered as poor, grades 4-5 as good collaterals [27]. Recanalization rate was assessed on digital subtraction angiography (DSA) at the end of endovascular treatment using the modified Treatment In Cerebral Ischemia (mTICI) scale. Patients with mTICI score of 2b-3 were considered as successfully recanalized, whereas patients with mTICI score ranging from 0 to 2a were classified as not [28]. CTP study was processed by commercially available delay-insensitive deconvolution automated software (Olea Sphere Version 3.0 SP23; Olea Medical, La Ciotat, France), using standard singular value decomposition method according to manufacturer instructions. All steps, including motion correction, smoothing and evaluation of time density curves, selection of arterial input and venous output functions, were checked for errors. As recommended by the vendor, total hypoperfused tissue and ischemic core volumes were defined as ischemic brain regions with Tmax threshold values >6 seconds (Tmax>6 seconds) and relative cerebral blood flow threshold values less than 40% of normally perfused tissue (rCBF<40%), respectively. The difference between Tmax>6 seconds lesion extent and rCBF<40% lesion size was considered as ischemic penumbra. Mismatch ratio was defined as the Tmax >6-second volumes divided by the rCBF<40% volume. Hypoperfusion Intensity Ratio (HIR) was defined as the ratio between Tmax >10-second lesion and Tmax >6-second lesion volumes, with HIR ≥0.4 predicting poor tissue-related collateral flow [29]. All these parameters were automatically segmented and calculated by the software. FIV was manually traced using ITK-SNAP software on follow-up NCCT at 24 hours after symptom onset/last known well blinded to the initial CTP data, EVT result and clinical outcome. FIV was automatically calculated by this software with a multislice planimetric method by summation of the hypodense areas, manually traced on each slice in which they were detectable, multiplied by slice thickness [30]. The presence of CTP infarct core at presentation in absence of FIV on 24 hours NCCT follow-up imaging was considered as absolute GIC (aGIC).

### Statistical Analysis

Categorical data were presented as absolute number (%), whereas continuous variables were summarized as mean ± Standard Deviation (SD) if normally distributed or median and interquartile ranges (IQR) in case of non-parametric distribution. Baseline and treatment characteristics and clinical outcomes were compared between patients with and without aGIC using a two-tailed, independent samples student t-test or Mann Whitney U test as appropriate according to the data distribution for continuous variables. Dichotomous variables were compared using the chi squared test. aGIC was the outcome of interest and its predictors were explored with logistic regression with backward elimination at p values <0.1. Potential predictors were selected based on univariate analysis and previous literature. aGIC prediction model was based on clinical and imaging variables readily available on admission, in the hyperacute phase of stroke assessment. Furthermore, variables for which associations were seen in univariable analysis were included as adjustment factors as well. The analyses were performed considering aGIC as a dependent variable. Statistical analyses were performed using the software package IBM SPSS Statistics version 23. A p value < 0.05 was considered statistically significant.

## RESULTS

A total of 652 patients undergoing the multimodal CT study protocol, successful EVT and an NCCT at 24 hours follow-up imaging were included in the analysis, as shown in Figure 1. The characteristics of the study population are summarized in Table 1. Thirty-five of 652 (5.3%) had aGIC. The median CTP aGIC volume was 26.2 mL (IQR 19.9-32.2). In the early time window, aGIC was seen in 32/394 (8.1%) patients. In the late time window, aGIC was found in 3/258 (1%) patients. In our cohort full recanalization (mTICI 2b-3) was present in 535/652 (82%) patients, with a not significant prevalence in aGIC than in non-aGIC group. Patients with aGIC had better ASPECTS at admission, shorter stroke onset-to-imaging time, poorer collateral status (P<0.001) and better functional outcome (P=0.010). CTP Infarct core size and penumbra volumes at admission did not differ significantly between these two groups. A significant (P=0.010) shorter onset-to-reperfusion time was present in aGIC group (P=0.002). As was predictable, since aGIC patients were defined by the absence of final infarct lesion at follow-up imaging, the infarct size difference in those groups was consequently significant, as well as NIHSS at discharge (P<0.001). In the adjusted analysis, variables that significantly differed between patients with and without aGIC were included in a binary logistic regression model with aGIC as a dependent variable. Multivariable logistic regression analysis showed that ASPECTS score (odds ratio [OR], 1.87; p<0.001), onset-to-imaging time (OR, 0.99; p=0.013), collateral score (OR, 0.45; p<0.004) and hypoperfusion intensity ratio (OR, 23.2; p<0.001) were independent predictors of aGIC after adjusting for potential confounders (Table 2). The median mRS at three-months was 2 (IQR 1-3) in patients with aGIC and 3 (IQR 1-5) in those without aGIC (P<0.05). Twenty-three of 35 (65.7%) patients with aGIC versus 270/617 (43.7%) patients without aGIC achieved a good outcome at three-months (P<0.05). Figure 2 shows an illustrative case of aGIC.

**Figure 1.**
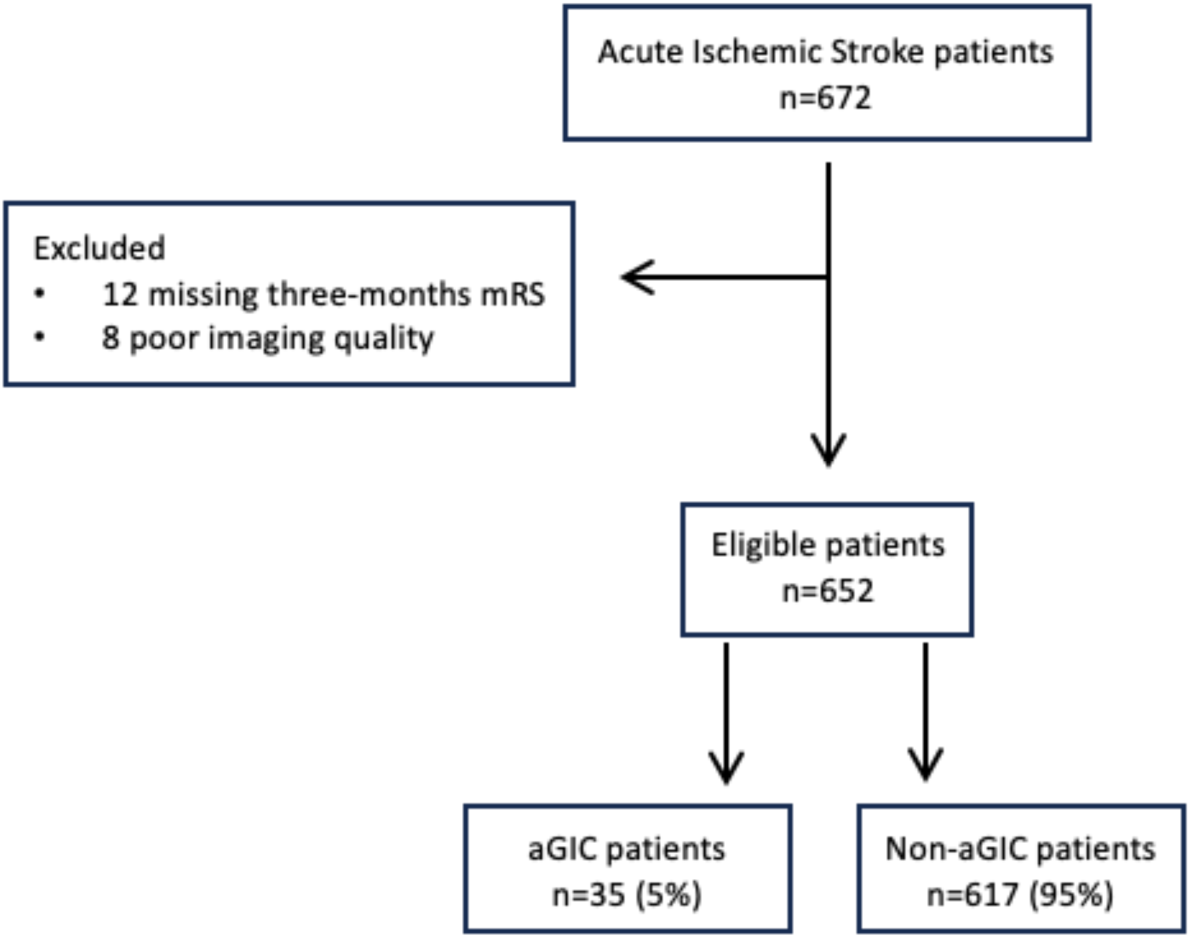
Flowchart of study population selection.

**Figure 2.**
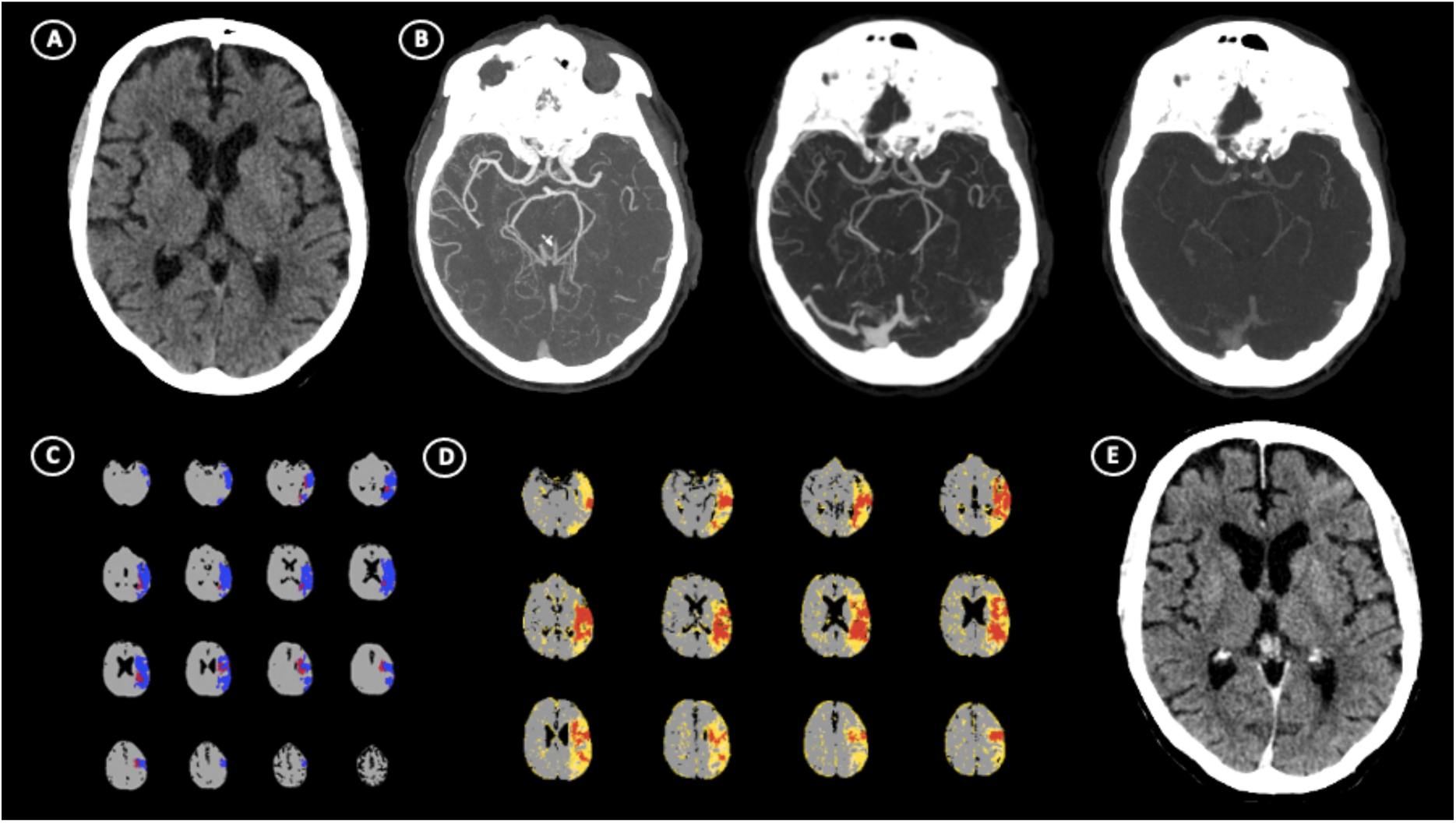
Illustrative case of absolute ghost infarct core (aGIC). A patient in the range of 60-65 years with acute ischemic stroke (AIS) suffering from the occlusion of left M1 segment of middle cerebral artery (MCA) occurred within 3 hours from stroke onset. (A) No visible hypodensity on non-contrast computed tomography (NCCT) resulted in ASPECTS=10 with (B) poor multi-phase CT-angiography collaterals (Menon Score=2). CT-Perfusion shows the presence of 14.5 mL of core volume and 83.6 mL of penumbra volume in left hemisphere (C) and high Hypoperfusion Intensity Ratio (HIR)=0.7 (D). After successful recanalization, follow-up NCCT performed at 24 hours (E) revealed no evidence of hypoattenuated areas indicating the absence of final infarct volume.

**Table 1.** Clinical and neuroimaging characteristics of patient’s population.

| Variable | Patients with<br>absolute ghost core | Patients without<br>absolute ghost core |
| --- | --- | --- |
|  | n = 35 | n = 617 |
| Age, mean (±SD), years | 76.0 (12.3) | 74.5 (12.9) |
| Sex, woman, n (%) | 18 (51.4) | 330 (53.4) |
| Admission NIHSS, median (IQR) | 16 (9-21) | 18 (12-22) |
| ASPECTS score, median (IQR) | 9 (8-10) | 8 (7-9) |
| rtPA before EVT, n (%) | 16 (45.7) | 248 (40.1) |
| Time from onset to NCCT, minutes, median (IQR) | 160 (90-250) | 290 (192-555) |
| Time from onset to NCCT <6 h, n (%) | 32 (91.4) | 362 (58.6) |
| Time from onset to NCCT >6 h, n (%) | 3 (8.6) | 255 (41.4) |
| Occlusion site, left n (%) | 23 (65.7) | 333 (53.9) |
| M1 segment, n (%) | 19 (54.3) | 364 (58.9) |
| M2 segment, n (%) | 9 (25.7) | 113 (18.4) |
| ICA segment, n (%) | 7 (20.0) | 140 (22.7) |
| Collateral Score, median (IQR) | 3 (3-4) | 4 (3-4) |
| Poor, n (%) | 23 (71.4) | 227 (36.7) |
| Good, n (%) | 12 (28.6) | 390 (63.3) |
| Hypoperfusion Intensity Ratio, median (IQR) | 0.50 (0.30-0.60) | 0.34 (0.20-0.50) |
| Infarct core (rCBF<40%) at baseline in mL, median (IQR) | 26.2 (19.9-32.2) | 23.7 (9.9-44.3) |
| Ischemic penumbra volume, median (IQR), mL | 65.0 (39.0-109.3) | 56.4 (28.5-93.1) |
| mTICI score 2b-3, n (%) | 35 (100.0) | 500 (81.0) |
| Onset to reperfusion time, minutes, median (IQR) | 330 (265-385) | 425 (310-715) |
| Infarct volume at 24h in mL, median (IQR) | 0 (0-0) | 29.5 (15.3-69.6) |
| NIHSS at 24h, median (IQR) | 4 (2-8) | 13 (5-20) |
| mRS at three-months 0-2, n (%) | 23 (65.7) | 270 (43.7) |

SD indicates Standard Deviation, NIHSS, National Institutes Health Stroke Scale; IQR, Interquartile Range; ASPECTS, Alberta Stroke Program Early CT Score; rtPA, tissue Plasminogen Activator; EVT, Endovascular Treatment; NCCT, Non-Contrast Computed Tomography; ICA, Internal Carotid Artery, rCBF, relative Cerebral Blood Flow; mTICI, modified Thrombolysis In Cerebral Infarction; mRS, modified Rankin Scale.

**Table 2.** Predictors of absolute ghost infarct core.

|  | Univariable |  | Multivariable |  |
| --- | --- | --- | --- | --- |
|  | OR (95% CI) | p | OR (95% CI) | p |
| Age, years | 1.02 (0.98-1.07) | .222 |  |  |
| Sex, woman | 0.82 (0.29-2.33) | .714 |  |  |
| Admission NIHSS | 1.05 (0.97-1.14) | .192 |  |  |
| ASPECTS score | 2.36 (1.52-3.67)) | <0.001 | 2.37 (1.61-3.48) | <0.001 |
| tPA before EVT | 0.67 (0.25-1.77) | .426 |  |  |
| Onset-to-CT time | 0.99 (0.98-0.99) | .016 | 0.99 (0.99-1.01) | .034 |
| Onset-to-reperfusion time | 1.00 (0.99-1.01) | .116 |  |  |
| mTICI score 2b-3 | 0.86 (0.31-2.56) | .996 |  |  |
| Collateral Score | 0.21 (0.10-0.42) | <0.001 | 0.24 (0.12-0.45) | <0.001 |
| Hypoperfusion Intensity Ratio | 8.88 (2.9-27.0) | <0.001 | 27.5 (1.54-48.9) | <0.001 |
| Infarct core (rCBF<40%) | 0.97 (0.95-1.00) | .010 |  |  |
| Ischemic penumbra volume | 0.99 (0.98-1.01) | .365 |  |  |
| NIHSS at 24 hours | 0.80 (0.72-0.89) | <0.001 |  |  |
| mRS at three-months 0-2 | 1.11 (0.38-3.25) | .023 |  |  |

NIHSS indicates National Institutes Health Stroke Scale; ASPECTS, Alberta Stroke Programme Early CT Score; tPA, tissue Plasminogen Activator; EVT, Endovascular treatment; mTICI, modified Thrombolysis In Cerebral Infarction.

## DISCUSSION

In this study, we defined ghost infarct core (GIC) as absolute GIC (aGIC) corresponding to the absence of final infarct lesion on 24 hours NCCT follow-up imaging in presence of CTP infarct core at presentation. In fact, as it is well-known that infarct can expand over time up to 5 days in both non-recanalized and recanalized subjects [21,22], ischemic volume assessed at 24 hours may underestimate final infarct lesion in patients achieving full recanalization [23,24]. Although edema formation undoubtedly contributes to infarct growth in early [19] and, mainly, in delayed [20] phases of ischemic evolution leading to a FIV overestimation, ischemic lesions increase after successful recanalization regardless of edema development [21]. Therefore, no infarct lesion at 24 hours associated with infarct core visible on admission CTP, as indicated by aGIC, seems to be a more reliable method for establishing the presence of GIC since no patients without infarcted tissue at 24 hours developed delayed FIV [23]. This approach substantially differed from all other studies defining GIC as an increase of 10 mL from the infarct core volume calculated on admission CTP within 24-48 hours [9–16] or up to 7 days [17]. As a consequence, in our study we found that the incidence of absolute ghost infarct core in recanalized patients was about of 5%, which was lower than in the majority of previous studies showing a frequency varying from 8.3% to 58.4% [9–15,17]. Of note, the frequency of GIC, defined as >10 ml of core size on admission CTP compared to the FIV calculated on 24 hours NCCT follow-up, was of 16.2% (106/652) in our population. In this regard, the most relevant difference was observed with studies adopting admission CBV maps to measure the infarct core size, in which ghost infarct core ranged from 38% to 58% of cases [9,11]. Taken together, these results suggest that CBV more commonly overestimates FIV than CBF maps confirming the higher predictive ability of CBF in correctly identifying infarct core compared to CBV [31]. Conversely, although the definition of ghost infarct core was different, the prevalence of this condition obtained in ours and in a recent study [16] was similar. A possible explanation of these findings is that, in the investigation of Sarraj and associates, median FIV size at 24-48 hours was of 0.8 mL, very close to the zero value that we used for classifying ghost infarct core in presence of an infarct lesion at admission CTP. Consistent with all previous studies [9–17], we found that ghost infarct core, as defined by aGIC, was significantly associated with a shorter time between symptom onset and the admission CTP, clearly prevailing in ultra-early presenters within 3 hours from last known well. It is not surprisingly that aGIC was a time-dependent phenomenon since brain tissue tolerance to ischemic insult depends not only on the severity, but also on the duration of ischemia [8,13,15], whereas decreased CBF values represent only the intensity, but not the duration of hypoperfusion [8,13,16]. Therefore, as CTP is able to describe hemodynamic changes but not the viability of brain tissue [16,17], a short ischemia duration can allow neuronal cells to tolerate very low CBF levels without development of cell death, leading to an infarct core overestimation by admission CTP [13]. In fact, lower CBF thresholds for better identifying infarct core were recently proposed in patients admitted early after symptom onset [16,32]. It is well-known that the presence of poor collateral extent assessed with single-phase CTA (sCTA) and/or HIR is the main factor determining a larger infarct core at admission that is overestimated when onset-to-imaging time is short [12,15–17]. Accordingly, we demonstrated that poor collaterals, as evaluated with mCTA and HIR, were significantly more represented in aGIC than in non-aGIC patients and were independently associated with aGIC. Compared to previous studies, our results appear more robust since obtained using both HIR for tissue-level and mCTA for pial arterial collateral measurements. In fact, while collaterals were graded with HIR in some publications [12,17], in other investigations they were scored with sCTA [15] or with HIR and sCTA [16]. However, it is currently accepted that mCTA is the more reliable tool for establishing pial arterial collateral filling [33]. Collateral assessment with mCTA was used only in the study of Ospel and coworkers [13] in which ghost infarct core was associated with good and not with poor collaterals probably because, unlike Menon score [27], moderate collaterals were considered as good. Finally, in line with three prior publications [12,13,15], in our study aGIC was associated with good functional outcome. This was an expected finding due to no evidence of FIV at 24 hours from onset characterizing our population. Nevertheless, the relationship between ghost infarct core and FIV was not reported in another study [17], confirming that FIV does not necessarily correlate to clinical outcome [34]. Our study has some limitations. First, this was a retrospective, single center analysis requiring a prospective validation. Second, the non-randomized design of our study introduced potential bias by unmeasured confounders. Third, generalizability might be limited as our analysis was restricted to patients selected for EVT. Finally, as recommended by the vendor, admission infarct core volume was calculated using rCBF<40% as threshold value instead of rCBF<30% that is widely considered the reference threshold value. Therefore, a confirmation of our findings using other platforms is needed. In addition, we did not perform a subgroup analysis with rCBF<20% to verify whether the incidence of ghost infarct core was limited by the use of lower CBF thresholds as previously suggested [16,17] since the corresponding CBF threshold for Olea software have not yet been identified.

## CONCLUSIONS

Ghost infarct core, as indicated by aGIC, is an uncommon phenomenon in ultra-early presenters associated with poor collaterals imaged within 3 hours after onset. The use of an absolute definition of GIC might provide a more reliable assessment of infarct core volume overestimation observed on admission CTP.

## Data Availability

Requests to access the dataset from qualified researchers trained in human subject confidentiality protocols may be sent to the corresponding author.

## ACKNOWLEDGEMENTS

None.

## DECLARATION OF CONFLICTING INTERESTS

The author(s) declared the following potential conflicts of interest with respect to the research, authorship, and/or publication of this article: Dr. Morotti declared consulting and expert meeting honoraria for EMG-REG International and AstraZeneca. Dr. Zini declared consulting and speaker fees from Boehringer-Ingelheim, Alexion-AstraZeneca and CSL Behring, Bayer, Angels Iniziative, Daiichi-Sankio. All the other Authors report no disclosures.

## FUNDING

The author(s) received no financial support for the research, authorship, and/or publication of this article.

## ETHICAL APPROVAL

The study has obtained approval from the Ethical Committee of the University of Firenze (PN 26299).

## INFORMED CONSENT

Written informed consent was obtained by patients or caregivers, or waived by the Institutional Review Boards.

## GUARANTOR

GB.

## CONTRIBUTORSHIP

GB and EF researched literature and conceived the study. GB, AM and IC conducted data analysis. GB and AM wrote the first draft of the manuscript, while EF provided significant intellectual contributions while editing the paper. All authors reviewed and edited the manuscript and approved the final version of the manuscript.

## Notes

### Funding Statement

No Funding

### Author Declarations

The study has obtained approval from the Ethical Committee of the University of Firenze (PN 26299).

